# Neoadjuvant chemotherapy response and genetic susceptibility in recently parous women with breast cancer

**DOI:** 10.1101/2025.02.13.25322229

**Authors:** Saya Dennis, Takahiro Tsukioki, Masha Kocherginsky, Andrea Keya Qi, Sarah DeHorn, Michael Gurley, Erica Wrubel, Yuan Luo, Seema Ahsan Khan

## Abstract

**Introduction:** Women with recent parity are at increased short-term breast cancer (BC) risk and face a worse prognosis. The effect of parity on response to neoadjuvant chemotherapy (NAC) is unstudied, and the influence of inherited susceptibility on parity-related short-term risk remains unclear.

**Methods:** We conducted a retrospective case-cohort study among women aged ≤50 with non-metastatic BC diagnosed between 2010 and 2020 who underwent genetic testing and were treated at Northwestern Medicine. Associations between NAC response and recency of parity were evaluated using multivariate logistic regression, stratified by tumor biologic subtypes. Relationships between germline mutations, recency of parity, and BC were explored via multi-state modeling and linear regression.

**Results:** Among 1,080 eligible women, 231 received NAC. Treatment response was poorer in parous women with triple negative tumors compared to nullipara, regardless of the recency of parity (*P*<0.03). Among 122 women (11.3%) with detectable pathogenic mutations, adjusted analyses with both modeling approaches revealed no indications that *BRCA1/2* carriers had an increased hazard of BC diagnosis in the decade following recent parity, compared to nulliparous mutation carriers. For *BRCA2* and *PALB2* carriers, breast cancer diagnosis occurred less frequently in the post-partum intervals.

**Conclusion:** We observed a poor response to NAC in parous TNBC patients compared to nullipara; effects of immunotherapy-based regimens deserve evaluation in the context of parity. Post-partum BC occurrence is not increased in *BRCA1/2* carriers; effects of rarer susceptibility genes may differ. These important effects of parity on BC in young women and those at genetic risk warrant larger prospective studies.

## Introduction

Breast cancer is the most common cause of cancer-related morbidity and mortality for women worldwide, and is one of the few cancers with rising incidence and mortality rate among <u>younger</u> women, particularly those of childbearing age [1, 2]. Younger women diagnosed with breast cancer, (age 40 years or less), face a higher risk of metastasis, resulting in a mortality rate twice that of older patients. [3–5] These trends are known to be influenced by important life events such as pregnancy and childbirth. Previous cohort studies showed that early first childbirth (in the 20s or earlier) protects against breast cancer development in the long term [6], but the decade that follows childbirth is associated with increased risk of developing breast cancer [7, 8]. Structural changes in the mammary gland, related to abrupt changes in hormone levels in the body during and after pregnancy, and remodeling of the extracellular matrix during postpartum mammary gland involution, are thought to increase the risk of developing breast cancer. [9]

Breast cancer diagnosed in women within 10 years of parity (post-partum breast cancer, or PPBC) displays less favorable biology than breast cancer in nulliparous women [10]. This includes a higher proportion with axillary nodal metastasis, absence of hormone receptor (HR), HER2 overexpression, high Ki-67, and high grade. [10–13] Moreover, a meta-analysis reported that PPBC carries a significantly higher mortality risk compared to non-PPBC [7], which is not solely explained by poor prognostic indicators [14–16]. Resistance to therapy may be an added disadvantage, and can be assessed in patients receiving neoadjuvant therapy, since treatment response provides a robust indicator of tumor sensitivity [17]. While data exist regarding response to neoadjuvant treatment when breast cancer is diagnosed within one year of term pregnancy [18, 19], its continuing impact within the post-partum decade has not been studied. Further investigation on this interplay may allow optimization of treatment strategies for women who have experienced recent parity (*i.e.* within the previous decade).

Another important aspect of BC in young women has been introduced by advances in genetic research that have revealed pathogenic/likely pathogenic variants (*i.e.* mutations) in tumor suppressor genes that significantly impact the development of early-onset breast cancer [20]. Young women with high penetrance susceptibility variants who are planning conception face decisions regarding the feasibility and effectiveness of intense breast surveillance during pregnancy and lactation, and worry about the potential for breast cancer development during or shortly after a term pregnancy. Understanding how parity interacts with genetic susceptibility is therefore crucial for optimizing risk-management in this high-risk population. The effect of parity on breast cancer risk in women with *BRCA1* and *BRCA2* mutations appears to differ from that of the general population. For instance, early first pregnancy —typically protective in the general population—may not offer the same benefit to *BRCA1/2* mutation carriers. [21–23], but parity in general may provide some protective effects. [24] Only one study has specifically examined the impact of the interval since the most recent parity on breast cancer risk in this group, showing reduced odds for breast cancer in *BRCA1* mutation carriers 1-2 years after childbirth and no effects for *BRCA2* carriers. [25] Since data in the general population show that breast cancer risk peaks around five years after childbirth and can remain elevated for over a decade [8], longer post-partum intervals need to be examined. The influence of mutations in more recently identified susceptibility genes on breast cancer risk following parity is unknown.

Our study addresses two previously unexplored aspects of breast cancer in recently parous women compared to their nulliparous counterparts: (1) associations between recency of parity and the patient’s response to neoadjuvant chemotherapy in the context of their tumor biology, and (2) exploration of the associations between the presence of germline pathogenic/likely pathogenic variants (mutations) and timing of breast cancer occurrence in relation to the patient’s most recent parity. Our results emphasize the need for collection of most recent parity data in large registries, so that these questions can be addressed in appropriately sized sample sets.

## Methods

This study was approved by the Institutional Review Board (IRB) of Northwestern University under protocol number STU00214082. The study design, data collection, and analysis procedures have been implemented in accordance with the approved protocol.

### Study population

We queried the Northwestern Medicine Electronic Data Warehouse (NMEDW) to select female patients aged 18 to 50 years, with an ICD code of invasive breast cancer or DCIS treated between 2010 and 2020. We selected patients who had a genetic counseling note documenting genetic testing. We confirmed that our cohort’s racial and ethnic distribution was representative of the overall breast cancer patients in the NMEDW using a chi-squared test. Demographic data, initial provider notes from breast cancer consultation, genetic counselling notes, pathological tumor staging summary notes, and use of neoadjuvant therapy were acquired by querying the NMEDW. A rule-based text mining algorithm was developed to automatically obtain the structured variables to the best extent possible. (https://github.com/sayadennis/recent-parity/tree/main/02_process_notes) The list of variables mined in each category of notes are shown in Supplementary Table 1. To ensure the accuracy of our text-mining algorithm, we calculated the agreement score between the manually reviewed data and the text-mined data for a random subset of 150 patients. The deviation was calculated using Concordance Correlation Coefficient (CCC) for continuous variables, and accuracy scores for categorical variables. (Supplementary Table 2) The CCC measures agreement between observed and predicted values, accounting for both magnitude and variability, with values near 1 indicating strong agreement and values near 0 reflecting poor concordance. The accuracy score, on the other hand, can be interpreted as the proportion of correctly labeled samples. For patients who received NAC, ER/PR/HER2 status were manually reviewed to ensure correct stratification during statistical analysis. Some data, including information from scanned documents and manually entered semi-structured notes, could not be retrieved by text-mining and was manually reviewed and completed by trained clinical experts (TT, SD, EW, AQ).

### Statistical Analysis

Our cohort consists of both parous and nulliparous cases. We defined “parity category” as a variable that takes one of four categories: a) nulliparous, b) most recent parity <5 years ago, c) 5–10 years ago, and d) ≥10 years ago, relative to age at breast cancer diagnosis. The patient’s response to neoadjuvant chemotherapy (NAC) was measured using a scoring system for Residual Cancer Burden (RCB), which is validated to be highly indicative of long-term outcomes such as relapse-free survival. [17, 26, 27] RCB is categorized as RCB-0 (pathological complete response), RCB-I (minimal burden), RCB-II (moderate burden), and RCB-III (extensive burden). We used multivariate logistic regression to examine the association between NAC response and parity categories, with our outcome of interest as poor response to NAC, defined as RCB-II or RCB-III. Our predictors included the four-level parity category with the nulliparous group as the reference category, age at diagnosis, and family history of breast cancer in first- or second-degree relatives.

In this case-cohort study, we could not use occurrence of breast cancer as the event of interest. Therefore, we used age of breast cancer diagnosis in specific post-partum intervals as a surrogate for whether risk was increased or decreased in a specific interval. We examined the association between mutation status, recent parity and age at breast cancer diagnosis by fitting linear regression models with age at diagnosis as the outcome variable, and parity category, mutation status, their interactions, and family history of breast cancer as predictors. Only *BRCA1*/*2* were included here since there were few carriers of other susceptibility genes. We assessed potential collinearity between mutation status and family history using a contingency table and McNemar’s test. Moreover, simpler models with and without family history as predictors confirmed that coefficients were stable across models and collinearity did not compromise model stability. To explore the potential interactions between mutation status and parity category, we assessed differences in diagnosis age between *BRCA1/2* mutation carriers and non-carriers within each parity category, using estimated marginal means followed by pairwise comparisons. [28] Given the exploratory nature of our study, we applied a significance threshold of 0.1.

We also used multi-state models [29–31] to explore the relationships between mutation status, parity, and time to breast cancer diagnosis. These models enable the analysis of hazard ratios (HRs) for covariates across transitions between more than two states. Our model included three states: The first is the nulliparous state (S0). The second state is defined as having had the most recent parity prior to breast cancer diagnosis (S1). Breast cancer diagnosis marked the final state (S2) (Figure 4a). Transition times between states were measured in years, with all patients transitioning to a breast cancer diagnosis (S2) by age 50, thus eliminating censoring. We estimated HRs for transitions between these states using the Cox proportional hazards model, adjusting for age and family history. Since Cox proportional hazards models are robust against sparse features, we included genes *BRCA1, BRCA2*, *PALB2*, *CHEK2*, and *ATM*. Analyses were done using R package mstate. [32]

## Results

### Patient Characteristics

We identified 2,606 patients with breast cancer ICD codes between 2010-2020 at age ≤50. After filtering for those treated at our institution and with verified genetic counselling at Northwestern, we were left with 1,211 patients; we then excluded 130 women who presented with metastatic disease, leaving a final sample set of 1,080 patients (Figure 1a). The proportion of patients aged >40 who underwent genetic testing increased during the study period, reflecting the increasing use of genetic testing over this timeframe. (Supplementary Figure 1) The racial/ethnic distribution of our cohort was similar to that of all women presenting to our center with breast cancer at age ≤50 over the same time-frame (Chi-statistic=0.05; *P*=0.99; Supplementary Table 3).

**Figure 1.**
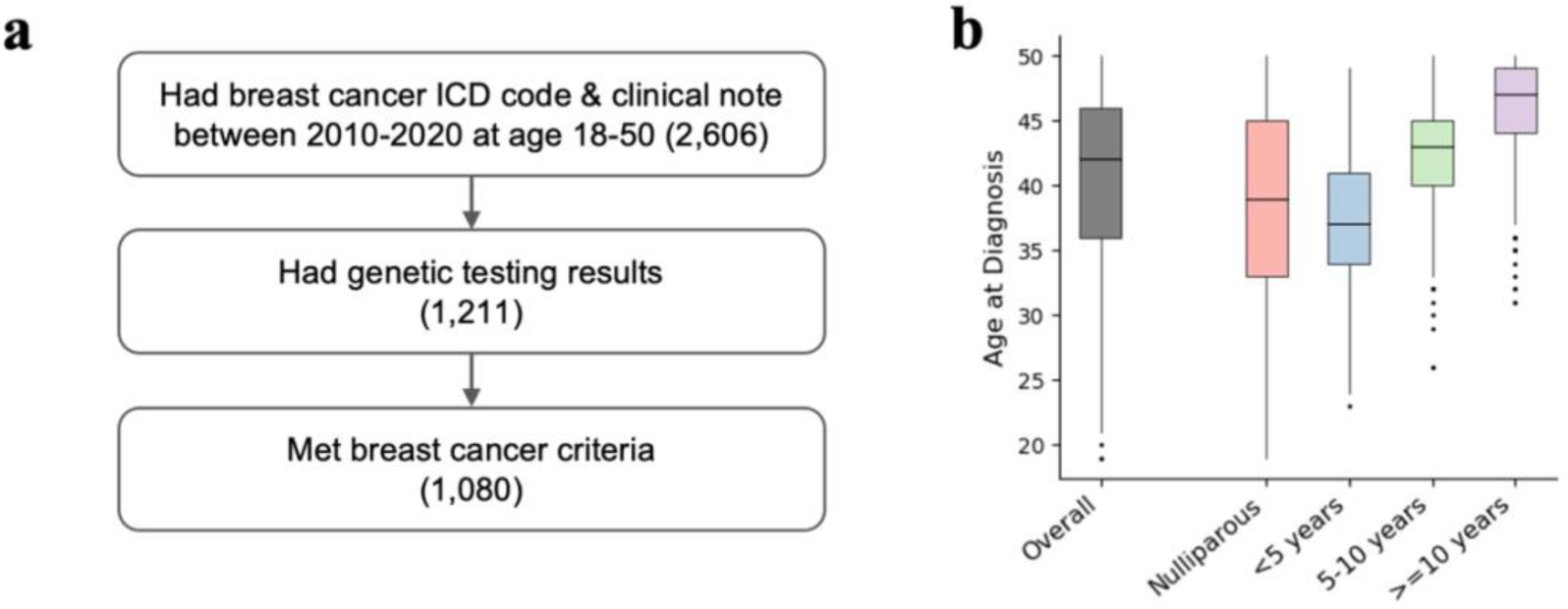
Summary of patient selection and distribution of breast cancer diagnosis age by parity category. **a)** Patient selection flowchart for narrowing down our cohort. Number in parentheses indicate the number of patients that remained after applying the given criteria. **b)** Distributions of the age of breast cancer diagnosis by parity category.

Table 1 summarizes our cohort’s demographic, parity history, and tumor characteristic data. We found that 421 women (38.9%) were nulliparous; 172 (15.9%) had breast cancer within 5 years of parity, 222 (20.4%) within 5-10 years, and 265 (24.6%) after ≥10 years. The proportions of patients with ER/PR+ HER2-, HER2+, and Triple Negative tumors were 66.0 %, 17.7 %, and 14.5 %, respectively. About 16.0 % of the patients had non-invasive (DCIS) histology.

**Table 1.** Summary of the demographic, gynecological, and tumor data.

|  | Overall<br>(N=1080) | Recency of Parity |  |  |  |
| --- | --- | --- | --- | --- | --- |
|  |  | Nulliparous<br>(n=421; 39%) | <5 years<br>(n=172; 16%) | 5-10 years<br>(n=222; 21%) | ≥10 years<br>(n=265; 24%) |
| Demographic |  |  |  |  |  |
| Mean Age at Dx<sup>1</sup> | 40.79 (±6.69) | 38.63 (±7.47) | 37.31 (±5.16) | 41.87 (±4.71) | 45.61 (±4.27) |
| Number of pregnancies<sup>1</sup> | 1.80 (±1.67) | 0.30 (±0.77) | 2.37 (±1.25) | 2.95 (±1.40) | 2.78 (±1.41) |
| Number of births<sup>1</sup> | 1.24 (±1.22) | 0.00 (±0.00) | 1.77 (±0.73) | 2.09 (±0.89) | 2.08 (±1.06) |
| Race/Ethnicity<sup>2</sup> |  |  |  |  |  |
| Asian | 85 (7.9%) | 31 (7.4%) | 18 (10.5%) | 17 (7.7%) | 19 (7.2%) |
| Black | 115 (10.6%) | 40 (9.5%) | 10 (5.8%) | 19 (8.6%) | 46 (17.4%) |
| Hispanic/Latinx | 110 (10.2%) | 30 (7.1%) | 19 (11.0%) | 24 (10.8%) | 37 (14.0%) |
| White | 720 (66.6%) | 302 (72.0%) | 115 (66.9%) | 152 (68.5%) | 150 (56.6%) |
| Other/ Unknown<sup>3</sup> | 49 (4.5%) | 16 (3.8%) | 10 (5.8%) | 10 (4.5%) | 13 (4.9%) |
| Breast cancer subtypes |  |  |  |  |  |
| ER/PR+, HER2- | 713 (66.0%) | 281 (66.7%) | 108 (62.8%) | 140 (63.1%) | 184 (69.4%) |
| HER2+ | 191 (17.7%) | 68 (16.2%) | 35 (20.3%) | 43 (19.4%) | 45 (17.0%) |
| Triple Negative | 157 (14.5%) | 66 (15.7%) | 25 (14.5%) | 35 (15.8%) | 31 (11.7%) |
| Missing | 19 (1.8%) | 6 (1.4%) | 4 (2.3%) | 4 (1.8%) | 5 (1.9%) |
| Histology |  |  |  |  |  |
| Non-invasive (DCIS) | 173 (16.0%) | 67 (15.9%) | 23 (13.4%) | 46 (20.7%) | 37 (14.0%) |
| Invasive | 861 (79.7%) | 336 (79.8%) | 139 (80.8%) | 165 (74.3%) | 221 (83.4%) |
| Other/Missing<sup>4</sup> | 46 (4.3 %) | 18 (4.3%) | 10 (5.8%) | 11 (5.0%) | 7 (2.6%) |
| Histologic grade |  |  |  |  |  |
| I | 289 (26.8%) | 116 (27.6%) | 43 (25.0%) | 62 (27.9%) | 68 (25.7%) |
| II | 340 (31.5%) | 140 (33.3%) | 62 (36.0%) | 59 (26.6%) | 79 (29.8%) |
| III | 142 (13.1%) | 53 (12.6%) | 27 (15.7%) | 25 (11.3%) | 37 (14.0%) |
| DCIS<sup>5</sup> | 173 (16.0%) | 67 (15.9%) | 23 (13.4%) | 46 (20.7%) | 37 (14.0%) |
| Missing | 136 (12.6%) | 45 (10.7%) | 17 (9.9%) | 30 (13.5%) | 44 (16.6%) |

<sup>1</sup> For these variables, the group's mean values (± standard deviations) are shown.
<sup>2</sup> Patients who are Hispanic and White or Hispanic and Black were categorized as Hispanic/Latinx.
<sup>3</sup> Some patients declined to provide their race or ethnicity information.
<sup>4</sup> Less than 1% of patients were missing histology for any parity category.
<sup>5</sup> DCIS only has nuclear grade and were therefore treated separately for the histologic grade counts. Women with DCIS were included in the analysis of genetic susceptibility and parity/breast cancer diagnosis.

### Neoadjuvant Treatment

In total, 231 patients (21.3 %) underwent neoadjuvant chemotherapy (NAC) (Figure 2). By subtype, 11.5% of ER/PR+, HER2-patients, 40.0% of HER2+ patients, and 53.3% of Triple Negative (TN) patients underwent NAC. (Supplementary Figure 1.a) NAC was more common among nulliparous women (24.9%) and those with parity <5 years (24.7%), while lower in women with parity ≥10 years (16.0%). Other factors associated with NAC use included TN or HER2+ tumors, nodal involvement, or younger age of diagnosis, aligning with current treatment standards. (Supplementary Figure 1.a-c) The number of patients receiving NAC increased over time, particularly for patients with HER2+ and TN tumors. (Supplementary Figure 1.d)

**Figure 2.**
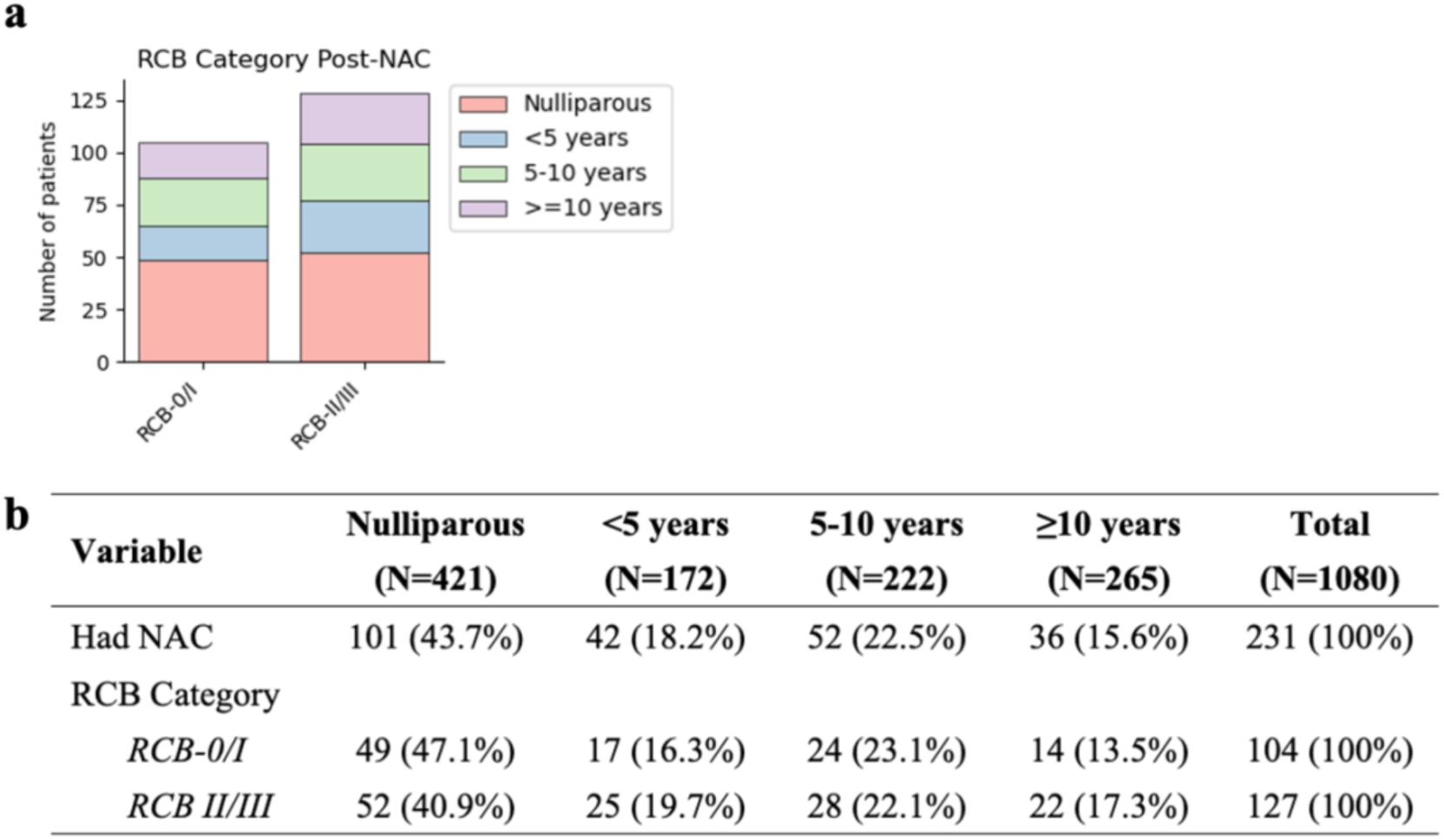
Summary of neoadjuvant chemotherapy (NAC) response. **a)** Graphical visualization of the number of patients that belonged to RCB-0/I or RCB-II/III, separated by parity category. (*RCB = Residual Cancer Burden); **b)** Table showing the number of patients who belonged to RCB-0/I or RCB-II/III.

We performed separate analyses by biologic subtype, (ER/PR+ HER2-, HER2+, and TN) due to their strong effects on NAC response. A chi-squared test indicated no significant association between clinical tumor stages and NAC response (χ²(3, N = 233) = 2.31, *P* = 0.67). We examined the association between parity status and poor NAC response (RCB-II or RCB-III), adjusting for age and family history. The results are shown in Table 2. In the TN group, parous women consistently demonstrated poorer responses to NAC (*i.e.* were more frequently classified as RCB-II or RCB-III) compared to nulliparous women, regardless of recency of parity. (OR=1.40, 1.47, and 1.54; *P*=0.0297, 0.0069, and 0.0324 respectively for <5 years, 5-10 years, and ≥10 years parity categories compared to nulliparous women.) This suggests that tumors developing in a background of parity may be less sensitive to regimens that were standard until 2020, even when parity is remote (≥10 years previously).

**Table 2.** Estimated odds ratio for poor response to neoadjuvant chemotherapy (RCB II-III) between women with parity compared to nulliparous women.

| Stratification | Outcome | Nulliparous | Parous <5 yrs ago |  |  | Parous 5-10 yrs ago |  |  | Parous ≥10 yrs ago |  |  |
| --- | --- | --- | --- | --- | --- | --- | --- | --- | --- | --- | --- |
|  |  | Counts (%) | Counts (%) | OR (90% CI) | P | Counts (%) | OR (90% CI) | P | Counts (%) | OR (90% CI) | P |
| ER/PR+, HER2- | RCB II-III | 30 (78.9%) | 9 (90.0%) | 1.10 (0.86-1.40) | 0.5304 | 10 (71.4%) | 0.93 (0.75-1.15) | 0.5733 | 13 (81.3%) | 0.99 (0.80-1.24) | 0.9698 |
|  | RCB 0-I | 8 (21.1%) | 1 (10.0%) |  |  | 4 (28.6%) |  |  | 3 (18.7%) |  |  |
| HER2+ | RCB II-III | 11 (40.7%) | 7 (41.2%) | 0.96 (0.75-1.24) | 0.8072 | 4 (23.5%) | 0.88 (0.67-1.16) | 0.4487 | 2 (20.0%) | 0.84 (0.60-1.17) | 0.3791 |
|  | RCB 0-I | 16 (59.3%) | 10 (58.8%) |  |  | 13 (76.5%) |  |  | 8 (80.0%) |  |  |
| Triple negative | RCB II-III | 9 (26.5%) | 9 (64.3%) | **1.40 (1.09-1.79)** | **0.0297** | 13 (68.4%) | **1.47 (1.17-1.85)** | **0.0069** | 7 (70.0%) | **1.54 (1.11-2.14)** | **0.0324** |
|  | RCB 0-I | 25 (73.5%) | 5 (35.7%) |  |  | 6 (31.6%) |  |  | 3 (30.0%) |  |  |

\*RCB = Residual Cancer Burden
\*Reference category for neoadjuvant chemotherapy response is RCB 0-I (good response). OR>1 indicates increased odds of poor response in a given parous category compared to nulliparous counterparts.
\*Estimates were obtained using separate logistic regression models for each biomarker category, adjusted for age at diagnosis and presence of family history.

### Genetic Susceptibility and effect of parity

The summary of our cohort’s genetic testing results is shown in Figure 3. Most patients (97.5%) had their genetic testing performed during or after their breast cancer diagnosis. Overall, pathogenic or likely pathogenic variants were found in 121 (11.2 %) women, and variants of uncertain significance (VUS) were detected in 96 (8.9 %). *BRCA1* and *BRCA2* had the highest counts of mutation carriers (58 and 38 respectively). There were few to no carriers of pathogenic variants for *PTEN*, *TP53*, *CDH1*, and *STK11*. Among *BRCA1* mutation carriers, 58.6% (34/58) were nulliparous, and among *BRCA2* carriers 42.1% (16/38) were nulliparous. (Figure 3b) The prevalence of family history of breast cancer was similar in the nulliparous and parous groups (46% and 45% respectively, Fisher’s exact *P* = 0.85),

**Figure 3.**
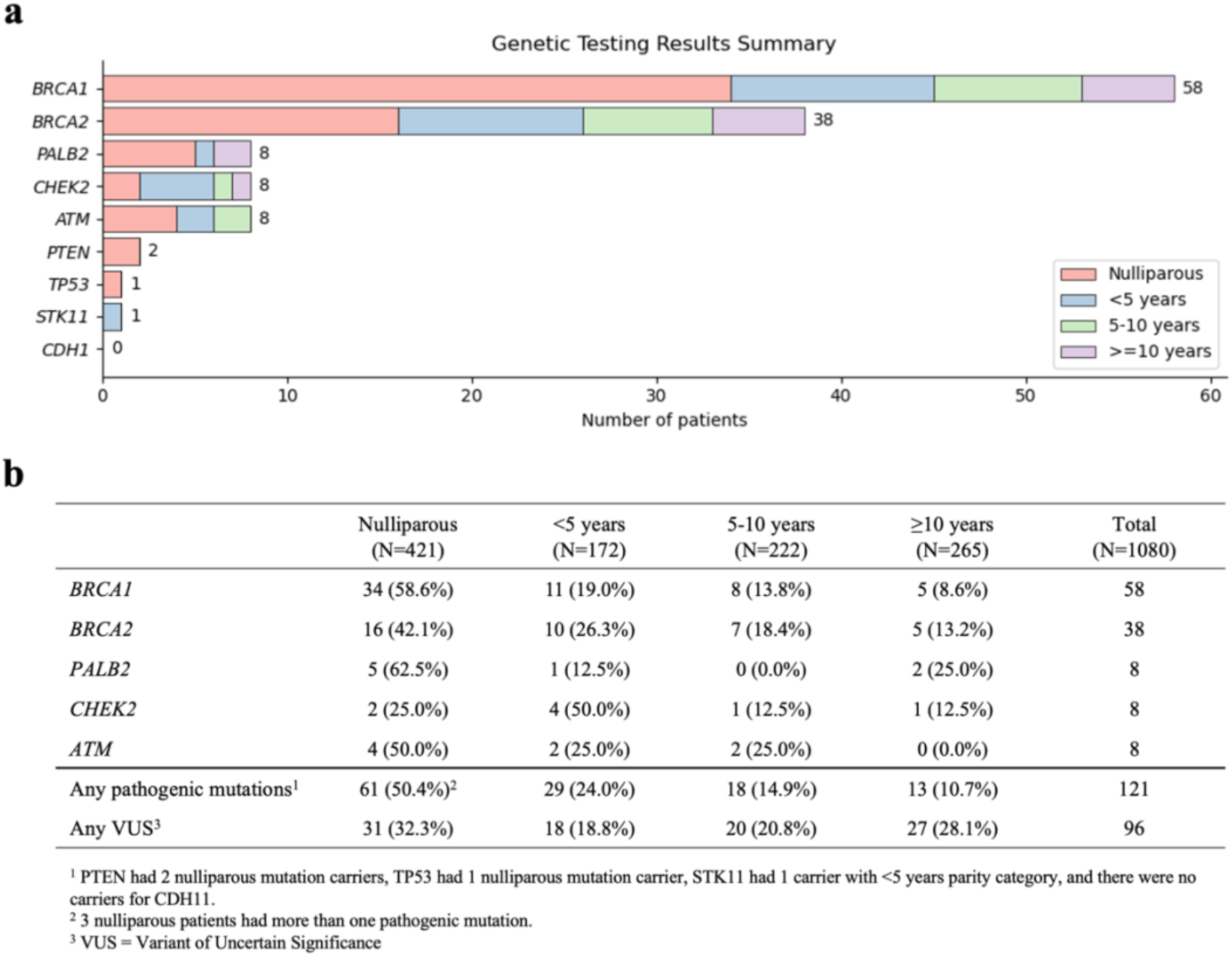
Summary of genetic testing results **a)** The distribution of parity categories of the patients who had pathogenic mutations in the specified genes. **b)** The number of pathogenic mutation carriers for each gene by parity category. Percentages are calculated as the proportion of mutation-carriers within each parity category.

The linear regression model allowed us to evaluate interactions between parity status and age of breast cancer occurrence among *BRCA1/2* carriers (there were too few carriers of other susceptibility genes for robust estimation using this method). We included four predictors: *BRCA1*/*2* mutation status, presence of family history, parity category, and interaction terms between mutation status and parity category. Collinearity between genetic mutation and family history was examined (Supplementary Figure 2), and was found to not obscure the model fit. Model diagnostics confirmed no major deviations from assumptions, ensuring the reliability of the results. (Supplementary Figure 3) Model estimates for the linear regression model are shown in Table 3. We first examined each parameter alone (*BRCA1*, *BRCA2*, and parity category). As expected, *BRCA1/2* mutation status were significantly associated with younger age at diagnosis compared to non-carriers. We then examined the interaction terms between parity category and *BRCA1/2* mutation status; these were not statistically significant overall. However, there was a trend in the interactions between *BRCA2* carrier status and parity <10 years before breast cancer diagnosis (unadjusted *P*<0.10); in this group the age of cancer diagnosis was delayed compared to nulliparous women of similar age, suggesting the possibility of a protective effect of parity in the decade following last parity. (Table 3) Supplementary Table 4 shows the estimated marginal means differences in age at breast cancer diagnosis for *BRCA1/2* mutation carriers compared to their non-carrier counterparts for each parity category. Nulliparous *BRCA1* carriers were diagnosed 3.63 years younger than their non-carrier counterparts (*P*=0.0024), and nulliparous *BRCA2* carriers were diagnosed 5.73 years younger than their non-carrier counterparts (*P*=0.0006).

**Table 3.** Estimated effects of mutation status and parity category on the linear regression model for age at cancer diagnosis. Negative regression coefficients (beta) indicate younger age at cancer diagnosis.

| Variable | Estimated effect (beta)<br>on Dx age in years<sup>2</sup> | 90% CI<sup>3</sup> | P-value |
| --- | --- | --- | --- |
| *BRCA1* (n=58)<sup>1</sup> | -3.6 | -5.4, -1.9 | <u><0.001</u> |
| *BRCA2* (n=38)<sup>1</sup> | -5.7 | -8.2, -3.3 | <u><0.001</u> |
| Has family history (n=491)<sup>1</sup> | 0.29 | -0.31, 0.88 | 0.4 |
| Parity category |  |  | <u><0.001</u> |
| Nulliparous | – | – |  |
| <5 years | -1.8 | -2.7, -0.85 | <u>0.002</u> |
| 5-10 years | 2.9 | 2.0, 3.7 | <u><0.001</u> |
| ≥10 years | 6.4 | 5.6, 7.2 | <u><0.001</u> |
| Interaction: *BRCA1* & Parity category |  |  | 0.4 |
| *BRCA1* & <5 years | 2.0 | -1.4, 5.4 | 0.3 |
| *BRCA1* & 5-10 years | -0.25 | -4.1, 3.7 | >0.9 |
| *BRCA1* & ≥10 years | 4.1 | -0.58, 8.8 | 0.15 |
| Interaction: *BRCA2* & Parity category |  |  | 0.14 |
| *BRCA2* & <5 years | 4.2 | 0.17, 8.2 | <u>0.086</u> |
| *BRCA2* & 5-10 years | 5.7 | 1.2, 10 | <u>0.035</u> |
| *BRCA2* & ≥10 years | 2.6 | -2.4, 7.7 | 0.4 |

\* Model fit with *BRCA1* and *BRCA2* mutation status, their interactions with parity category, adjusted for family history status (yes/no).
<sup>1</sup> The “n” indicates the number of patients who had positive value for the given binary variable.
<sup>2</sup> Estimated linear regression coefficients (Betas). Negative value indicates breast cancer diagnosis at a younger age.
<sup>3</sup> CI = Confidence Interval

Our multi-state model allowed us to evaluate five genes: *BRCA1*, *BRCA2*, *PALB2*, *CHEK2*, and *ATM* in relation to breast cancer onset in parous versus nulliparous states. We obtained estimates of hazard ratios (HRs) for the interval between age 18 and breast cancer diagnosis if they remained nulliparous (T_0,2_), or from most recent parity to breast cancer diagnosis (T_1,2_), by genetic mutation. (Figure 4a) HRs were adjusted for family history and age at most recent parity for parous women. (Figure 4a) Women with *BRCA1* mutations had higher breast cancer hazard regardless of parity status (HR=1.92, *P*=0.00028 and HR=1.55, *P*=0.030 respectively). Women with *BRCA2* mutations also had a higher hazard for nulliparity-to-BC (HR=1.75, *P*=0.032) but not for parity-to-BC (HR=1.40, *P*=0.12). *PALB2* showed high HR for nulliparity-to-BC (HR=3.82, *P*=0.0032), while *ATM* showed elevated HR for parity-to-BC (HR=2.55, *P*=0.063) (Figure 4b). Hazard ratios for these two transitions were not significantly from each other. (Supplementary Table 5)

**Figure 4.**
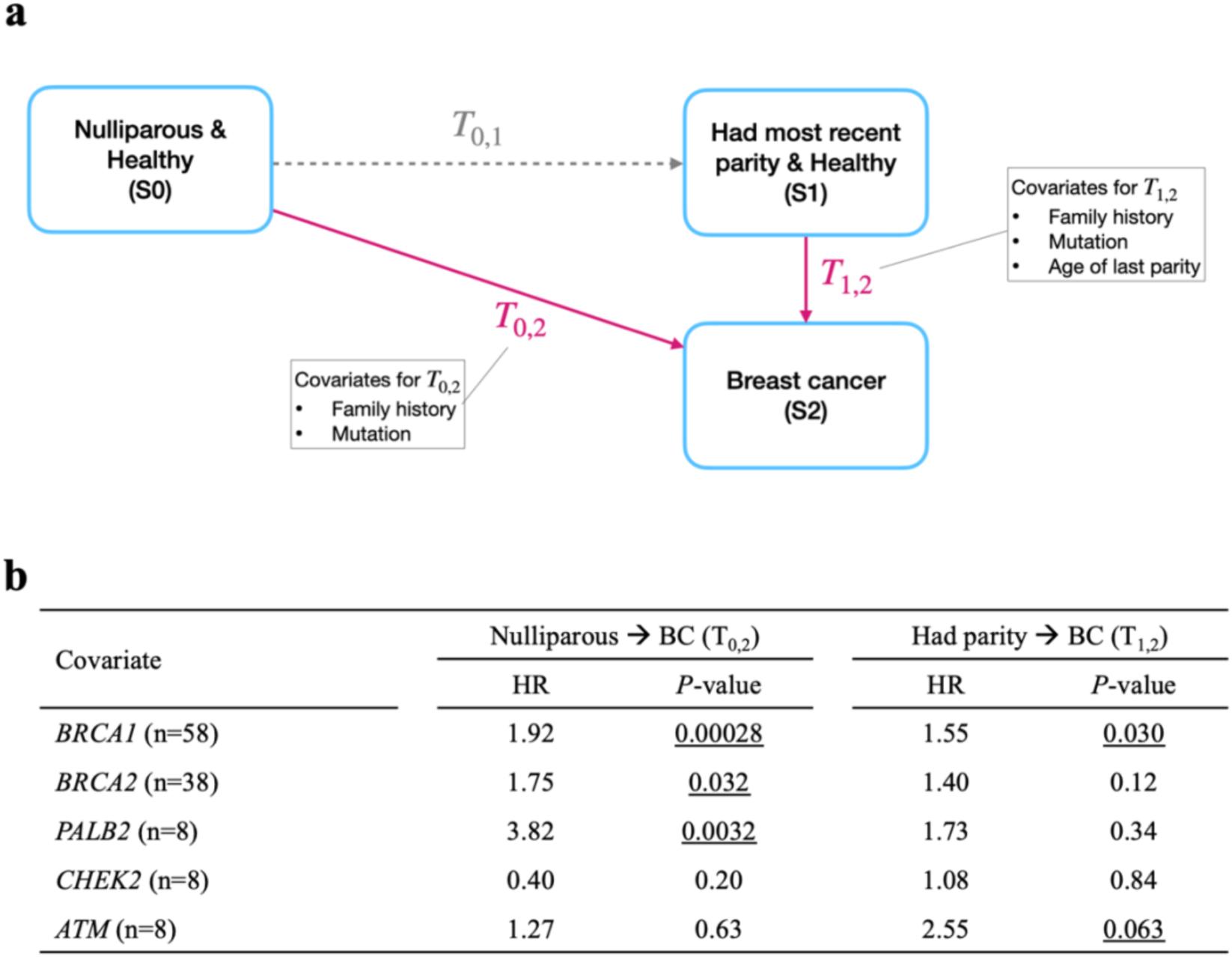
Results of multi-state modeling. **a)** Transition states diagram of the multi-state model. Our two transitions of interest are highlighted in magenta (T_0,2_ and T_1,2_). **b)** Hazard ratios (HRs) comparing pathogenic mutation carriers vs. non-carriers for our genes of interest, and their corresponding P-values, for transitions from nulliparity to cancer diagnosis and from last parity to cancer diagnosis. Sample size “n” indicates the number of patients with positive genetic testing results for that gene. A high HR indicates increased hazard of transition for mutation carriers compared to non-carrier counterparts.

## Discussion

We aimed to investigate two aspects of post-partum breast cancer: (1) associations between recency of parity and tumor response to neoadjuvant chemotherapy, and (2) exploratory associations of the timing of breast cancer in relation to the patient’s most recent parity in the presence of germline pathogenic mutations. Our findings shed light on important aspects of breast cancer management in young women and provide insights into areas that require further investigation.

Our analysis revealed poorer NAC response in parous women with triple-negative breast cancers (TNBC) with consistent enrichment in RCB-II/III categories across all parity intervals. The treatment of TNBC followed the standard-of-care regimen used until 2020, consisting of a sequential anthracycline-taxane based approach. Given the well-documented immune environment of TNBC, it is quite possible that the inclusion of immunotherapy agents will substantially address this difference in NAC response among parous and nulliparous women. Nevertheless, the potential role of parity in treatment response has not previously been reported and if confirmed in larger studies, deserves further investigation in terms of both the biology of breast cancers occurring in the post-partum interval and the selection of treatment strategies.

Our study also revealed intriguing results regarding the relationship between parity and germline pathogenic mutations. Given our case-cohort design, we used a linear regression model to evaluate interactions between parity category and the two most frequent gene mutations (*BRCA1/2*) with regards to age at breast cancer occurrence. We evaluated overall and each parity category to examine whether their combined presence hasten or delay the age at BC diagnosis compared to nulliparous women. In addition, the multi-state model investigated transitions to breast cancer from nulliparity and from most recent parity, with three additional genes (*PALB2, CHEK2, ATM*).

The linear regression analysis indicated a significantly younger diagnosis age overall for *BRCA1*/*2* carriers compared to non-carriers. We saw no significant interactions between mutation status and parity category with regards to breast cancer diagnosis age among parous carriers overall. However, *BRCA2* carriers within parity intervals of 0-5 and 5-10 years had a significant delay in diagnosis, which suggests a decreased likelihood of breast cancer development in the 10 years that follow childbirth (Table 3), possibly indicating that for *BRCA2* carriers, recent childbirth within these timeframes delays the age of breast cancer diagnosis compared to nulliparous women of similar age.

The multistate model allowed us to identify differences in transition to breast cancer between carriers and non-carriers in parous and nulliparous women. Our findings are reassuring to carriers of the most prevalent mutations of *BRCA1/BRCA2* in that the post-parity interval revealed no higher hazard for breast cancer diagnosis compared to nulliparous women of similar age. (Supplementary Table 3) However, we observed a higher hazard ratio (HR) for nulliparous *PALB2* carriers and parous *ATM* carriers in transition to BC diagnosis, although numbers were small. These hypothesis-generating findings suggest a link between non-*BRCA* mutations and parity in breast cancer development and points to the need for larger studies where date of last parity before breast cancer diagnosis is known.

We used two modeling approaches: linear regression and a multi-state model. Linear regression estimates how predictors affect the absolute age of breast cancer diagnosis, focusing on a static outcome—age at diagnosis. It can however capture non-linear relationships between childbirth timing and diagnosis age, important since prior data suggest that the effect of recent parity on breast cancer risk is non-linear. [8] In contrast, the multi-state model is dynamic, allows time-dependent effects, and is robust against sparse data due to its semi-parametric nature, enabling exploration of non-*BRCA* genes. The two models’ findings are consistent for *BRCA1*/*2* carriers. Linear regression showed similar diagnosis ages for parous *BRCA* carriers and non-carriers within each parity category (Table 3). The multistate model showed no excess risk in the post-partum interval compared to nullipara. However, linear regression results suggest childbirth may have a protective effect in *BRCA2* and *PALB2* mutation carriers, delaying diagnosis age compared to non-carriers. These findings need confirmation in larger studies.

Our findings on the relationship between recency of parity, genetic susceptibility, and breast cancer complement existing literature. Cullinane et al. found decreased odds of breast cancer diagnosis in the 1-2 years following parity in *BRCA1* carriers, while *BRCA2* carriers showed no significant trends. [25] Our results align with theirs, although our case-cohort design differs from their case-control design and the use of odds ratios. These findings should be extended to larger prospective cohorts that include affected and unaffected carriers with detailed parity history, including most recent parity age.

The main limitation of our study was an overall limited sample size and small numbers of non-*BRCA1/2* genes, and the case-cohort design which led us to use age of diagnosis as a surrogate for risk. Although our findings need confirmation in larger case-control cohorts, a general limitation in studies addressing post-partum BC risk is the lack of readily available data on recency of parity. While the patient’s age at first parity is routinely recorded, the age of the most recent childbirth prior to cancer is not often documented; preventing us from expanding our cohort to other institutions or large registry datasets. To advance our understanding in this area, it is essential to make systematic and routine efforts to record information about the time of most recent parity prior to cancer diagnosis. Finally, while sample size limited our ability to assess the connections between NAC response and genetic susceptibility, this remains a promising area for future exploration.

Overall, our results emphasize the need for further exploration in two important understudied aspects of breast cancer and parity history. The poorer prognosis of post-partum breast cancer is well documented, but the response to neoadjuvant therapy has not been previously examined. Additionally, while *BRCA1*/*2* mutations have long been the focus of research, the specific effects of the timing of recent parity needs further definition since it can impact important decisions regarding surveillance and mastectomy for women considering childbearing. These investigations should also be extended to include other cancer susceptibility genes. As more data accumulates on genetic testing on non-*BRCA1/2* genes, diversification of genetic research will be essential for data-based counselling of genetically susceptible women who are planning pregnancy, and appropriate therapeutic interventions for recently parous women.

## Data Availability

Deidentified data produced in the present study are available upon reasonable request to the authors

## Acknowledgements

We gratefully acknowledge the support from the Bluhm Family Foundation. This research was supported in part by the Cancer Center Support Grant (CCSG) P30 CA060553, and the NIH grant R01LM013337. Additionally, we acknowledge the resources and support provided by the Quantitative Data Sciences Core of the Lurie Cancer Center and the Northwestern Medicine Enterprise Data Warehouse.

## Supplementary Tables

**Supplementary Table 1.** List of collected variables and their sources.

| Source | Variable names |
| --- | --- |
| Initial counsel notes | • Age of menarche<br>• Gravida/para<br>• Age at first live birth<br>• Age at most recent live birth |
| Genetic counsel notes | Pathogenic mutations or variants of uncertain significance (VUS) in:<br>BRCA1, BRCA2, PALB2, TP53, CHEK2, PTEN, CDH1, STK11, ATM |
| Tumor staging summary notes | • Histology<br>• Tumor size<br>• Histologic Grade<br>• Tumor staging category<br>• Node staging category<br>• ER/PR status<br>• HER2 status |

**Supplementary Table 2.** Summary of the agreement between chart-reviewed data and text-mined data.

| Variable Name | Concordance Correlation Coefficient<br>(Continuous variables) | Accuracy Score<br>(Categorical variables) |
| --- | --- | --- |
| Age at diagnosis | 0.950 | - |
| Number of pregnancies | 0.965 | - |
| Number of births | 0.985 | - |
| Age at first pregnancy | 0.995 | - |
| Age at most recent pregnancy | 0.982 | - |
| Tumor size | 0.921 | - |
| Histology | - | 0.900 |
| Histologic grade | - | 0.930 |
| ER status | - | 0.989 |
| PR status | - | 0.966 |
| HER2 status | - | 0.930 |

**Supplementary Table 3.** Distributions of race and ethnicity in our cohort alongside those of all women diagnosed with breast cancer at age ≤50 between 2010 and 2020, regardless of whether they received genetic testing.

|  | Asian | Black | Hispanic/Latinx | White | Other/Unknown |
| --- | --- | --- | --- | --- | --- |
| Our cohort<br>(has genetic testing) | 7.9% | 10.6% | 10.2% | 66.6% | 4.5% |
| All women regardless of<br>presence of genetic testing | 5.6% | 9.5% | 7.8% | 66.2% | 10.9% |

**Supplementary Table 4.** Estimated differences in age at breast cancer diagnosis between *BRCA1/2* mutation carriers vs. non-carriers.

| Parity category | Comparisons<sup>1</sup> | Estimated difference<br>in years<sup>2</sup> | *P*-values<sup>3</sup> |
| --- | --- | --- | --- |
| Nulliparous | BRCA1: yes vs. no | **-3.63** | **0.0024** |
|  | BRCA2: yes vs. no | **-5.73** | **0.0006** |
| <5 years | BRCA1: yes vs. no | -1.63 | 0.4749 |
|  | BRCA2: yes vs. no | -1.54 | 0.5641 |
| 5-10 years | BRCA1: yes vs. no | -3.87 | 0.1359 |
|  | BRCA2: yes vs. no | -0.01 | 0.9960 |
| $\geq 10$ years | BRCA1: yes vs. no | 0.50 | 0.8514 |
|  | BRCA2: yes vs. no | -3.10 | 0.4882 |

<sup>1</sup> For a given contrast, estimated marginal means are reported based on a linear regression model, and are averaged across other predictors in the model. For example, the BRCA1 contrast for the nulliparous category is averaged over the levels of BRCA2 and family history status.
<sup>2</sup> Estimated contrasts in years. A negative value indicates mutation carriers are diagnosed at a younger age than non-carriers.
<sup>3</sup> A significant *P*-value indicates that age of diagnosis is significantly different between mutation carriers and non-carriers for that given parity category.

**Supplementary Table 5.**
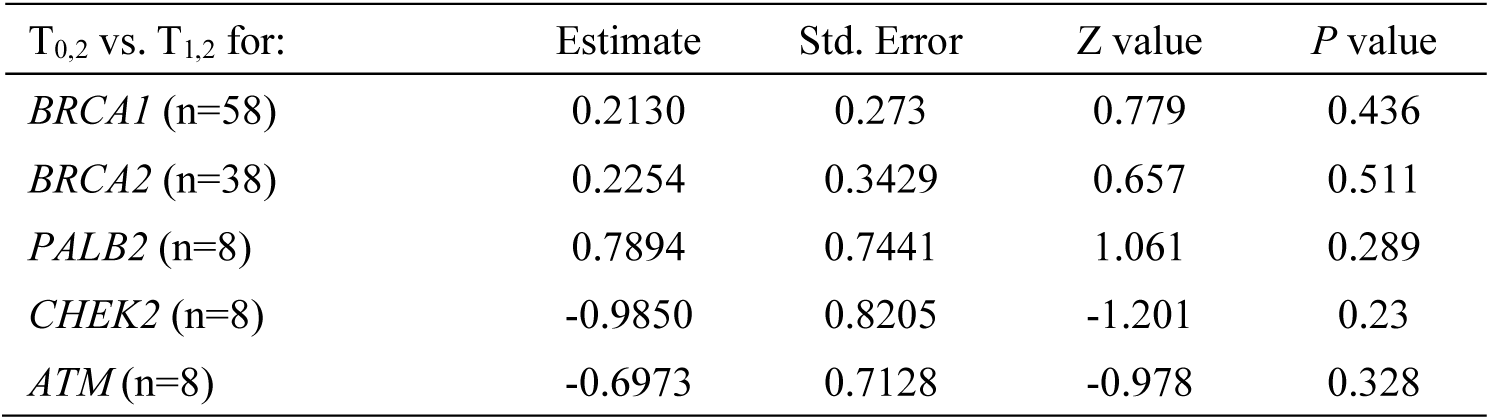
Results of pairwise comparisons of estimated coefficients using general linear hypothesis testing (GLHT) for multi-state modeling.

## Supplementary Figures

**Supplementary Figure 1.**
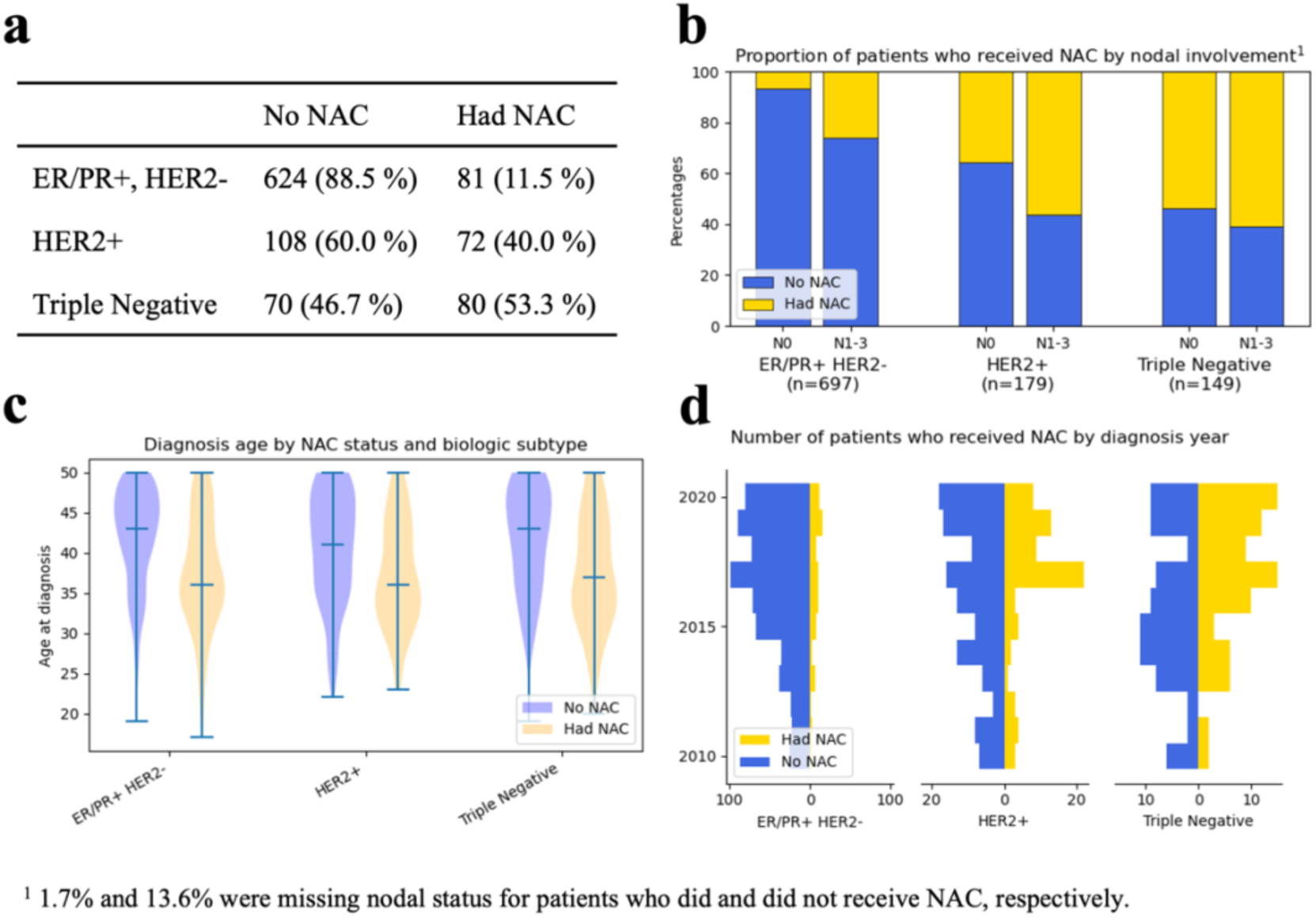
Differences between patients who did or did not receive neoadjuvant chemotherapy (NAC).

**Supplementary Figure 2.**
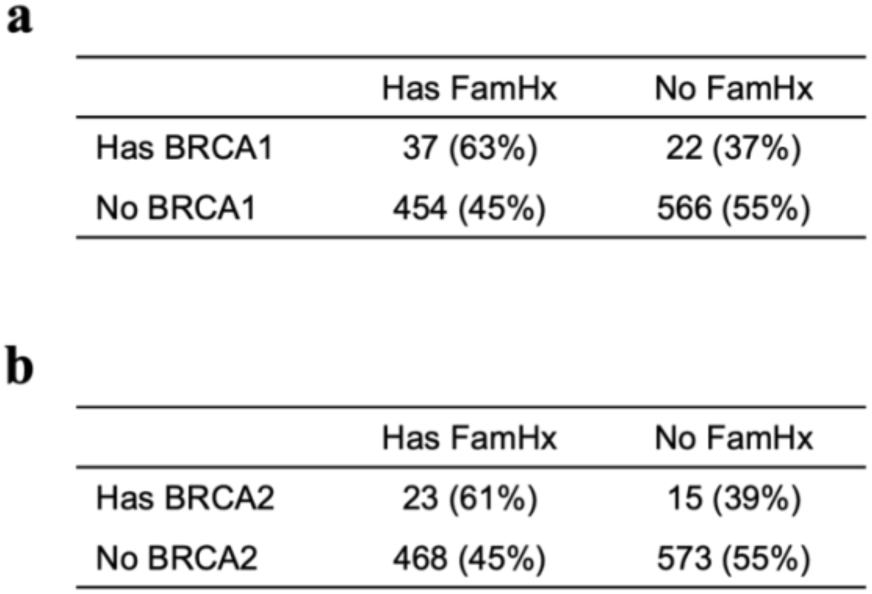
Contingency tables for the presence of family history (history of breast cancer in first of second degree relative) and BRCA1 (a) and BRCA2 (b) mutation status. McNemar’s test resulted in p < 2.2e-16 for both tables.

**Supplementary Figure 3.**
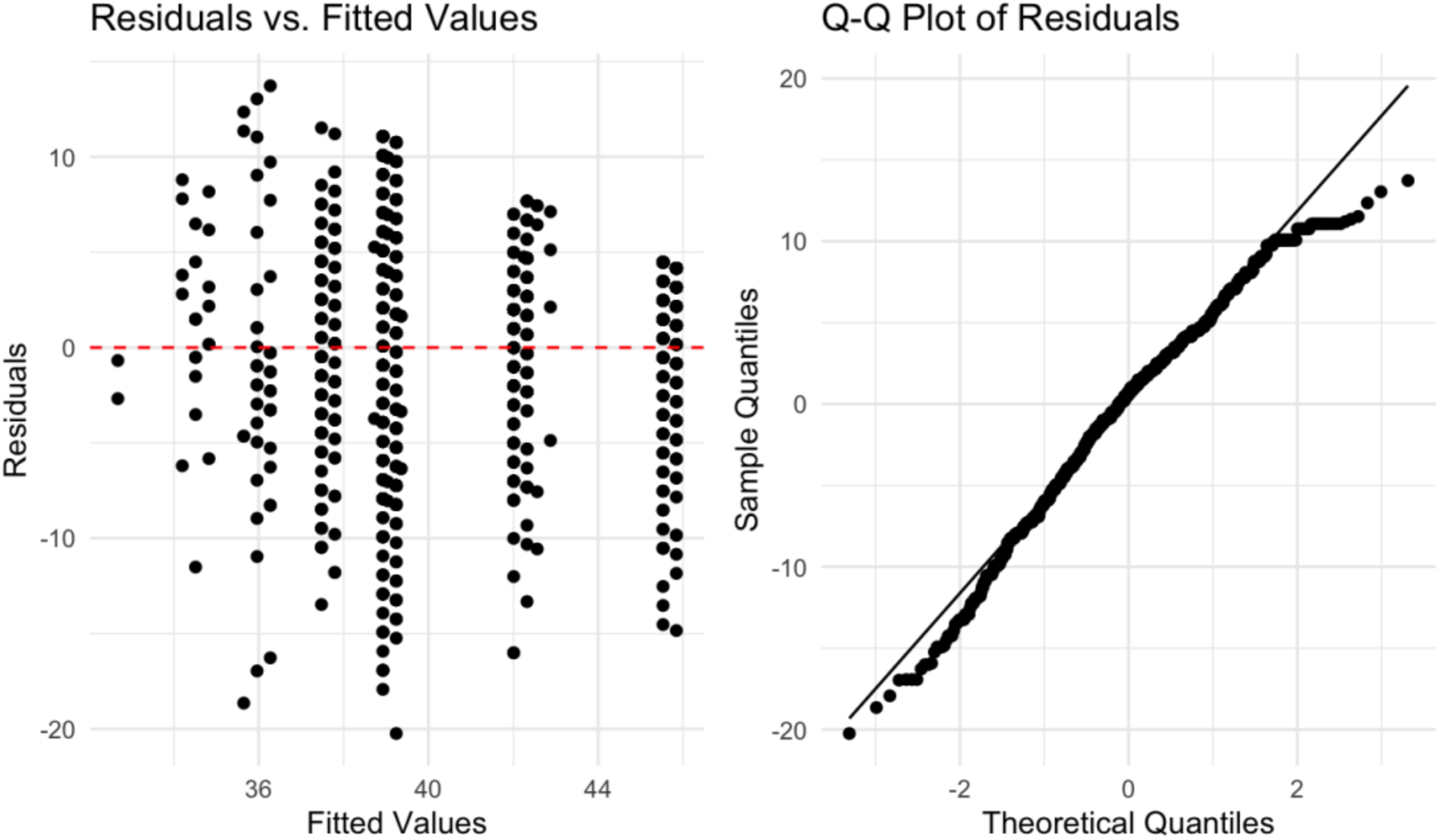
Residuals plot and Q-Q plot of the fitted linear regression model where age at diagnosis is the outcome variable, and parity category, mutation status, and family history are predictor variables.

## Notes

### Competing Interest Statement

The authors have declared no competing interest.

### Author Declarations

This study was approved by the Institutional Review Board (IRB) of Northwestern University under protocol number STU00214082.

